# Cardiac Adverse Events of Immune Checkpoint Inhibitors in Oncology Patients: A Systematic Review and Meta-Analysis

**DOI:** 10.1101/2020.06.01.20119719

**Authors:** Nso Nso Vialli, Daniel Antwi-Amoabeng, Bryce D. Beutler, Mark B. Ulanja, Jasmine Ghuman, Ahmed Hanfy, Joyce Nimo-Boampong, Sirri Atanga, Rajkumar Doshi, Sostanie Enoru, Nageshwara Gullapalli

## Abstract

**Background:** Immune checkpoint inhibitors (ICIs) are novel therapeutic agents used for various types of cancer. ICIs have revolutionized cancer treatment and improved clinical outcomes among cancer patients. However, immune-related adverse effects of ICI therapy are common. Cardiovascular immune-related adverse events (irAEs) are rare but potentially life-threatening complications. We aimed to estimate the incidence of cardiovascular irAEs among patients undergoing ICI therapy for various malignancies.

**Methods:** We conducted this systematic review and meta-analysis by searching PubMed, Cochrane CENTRAL, Web of Science, and SCOPUS databases for relevant interventional trials reporting cardiovascular irAEs. We performed a single-arm meta-analysis using OpenMeta [Analyst] software of the following outcomes: myocarditis, pericardial effusion, heart failure, cardiomyopathy, atrial fibrillation, myocardial infarction, and cardiac arrest. We assessed the heterogeneity using the I^2^ test and managed to solve it with Cochrane’s leave-one-out method. The risk of bias was performed with the Cochrane’s risk of bias tool.

**Results:** A total of 26 studies were included. The incidence of irAEs follows: myocarditis: 0.5% (95% CI [0.1%-0.9%]); pericardial effusion: 0.5% (95% CI [0.1%-1.0%]); heart failure: 0.3% (95% CI [0.0%-0.5%]); cardiomyopathy: 0.3% (95% CI [-0.1%-0.6%]); atrial fibrillation: 7.6% (95% CI [1.0%-14.1%]); myocardial infarction: 0.4% (95% CI [0.0%-0.7%]); and cardiac arrest: 0.4% (95% CI [0.1%-0.8%]).

**Conclusion:** The most common cardiovascular irAEs were atrial fibrillation, myocarditis, and pericardial effusion. Although rare, data from post market surveillance will provide estimates of the long-term prevalence and prognosis in patients with ICI-associated cardiovascular complications.

## Introduction

Immune checkpoint inhibitors (ICIs) have demonstrated remarkable efficacy in various malignancies, including lung cancer, melanoma, Hodgkin’s lymphoma, bladder cancer, and microsatellite instability (1). ICIs exert their effects through blocking inhibitory receptors on tumor cells (programmed cell death 1 ligand-1 [PD-L1]) (2, 3) or T-lymphocytes (programmed cell death protein-1 [PD-1] or cytotoxic T lymphocyte-associated protein-4 [CTLA-4]) (4, 5). The blockade of these receptors activates the effector T cells to target neoplastic cells (2). Many studies have demonstrated significant survival benefits of ICIs (6–8) and over 1,200 trials are currently ongoing (9).

The mechanism of action of ICIs involves non-specific activation of the immune system (10). Consequently, autoimmune inflammatory reactions frequently occur; this can ultimately lead to a broad spectrum of immune-related adverse events (irAEs) affecting both on-target and off-target organs (11). Reactions involving the skin, gastrointestinal tract, and endocrine system are relatively common among cancer patients on ICIs (12, 13). Approximately 80% of patients treated with agents targeting CTLA-4, 70% of patients treated with anti-PD-1 drugs, and 40% of those treated with anti-PD-L1 agents develop irAEs (13, 14). Severe events are common and up to 40% of patients on ICIs require treatment discontinuation due to irAEs (10).

Cardiovascular irAEs are rare, but potentially life-threatening (15). Although the initial trials on ICIs did not assess myocardial activity, growing evidence from case reports, case series, and cohort studies have raised awareness of unexpected cardiac toxicities associated with ICI therapy (16–18). Dual therapy appears to markedly increase the risk of cardiovascular irAEs; using the Bristol-Myers Squibb safety database, the estimated rate of myocarditis in patients receiving combination immunotherapy (ipilimumab plus nivolumab) was 0.27% as compared to 0.06% in those receiving nivolumab monotherapy (18).

Data on other ICI-related cardiac toxicities are scarce. This study aims to provide high-class evidence on the incidence of ICI-related cardiovascular adverse events through a systematic review and meta-analysis.

## Methods

This systematic review and meta-analysis complies with the Preferred Reporting Items for Systematic reviews and Meta-Analyses (PRISMA) statement (19) and Cochrane’s Handbook of Systematic Reviews of Interventions (20).

### Eligibility criteria

Our analysis included interventional trials involving patients receiving an ICI in which an adverse cardiovascular event was reported. We excluded the following: non-randomized trials, trials involving concurrent use of other anticancer interventions, animal studies, non-clinical studies, reviews, and meta-analyses. We also excluded studies without accessible data, conference abstracts, and studies for which there was no English language translation.

### Literature search

We searched PubMed, Cochrane CENTRAL, SCOPUS, and Web of Science databases for possible included articles according to our eligibility criteria from May 1^st^, 2020 through May 15^th^, 2020. We retrieved articles using a combination of the following keywords: “cardiotoxicity”, “adverse”, “events”, “myocard*”, “pericard*”, “neoplasm”, “cancer”, and “immune checkpoint inhibitor.”

### Study Selection, Data collection, and analysis

#### Screening of results

We performed the screening of retrieved studies through two stages. The first stage involved the inclusion and exclusion of studies based on title and abstract review. Selected studies underwent full-text screening against the inclusion criteria. Studies that had a mismatch with a single inclusion criterion were excluded. We conducted another search through the references of the included trials to ensure that no trials were inadvertently excluded. We considered studies which included multiple treatment arms as separate studies based on the adverse event reporting and refer to them as first author last name, year of publication followed by a, b, or c in the forest plot diagrams and Table 1. Figure 1 shows a PRISMA flow chart of the literature search.

**Figure 1.**
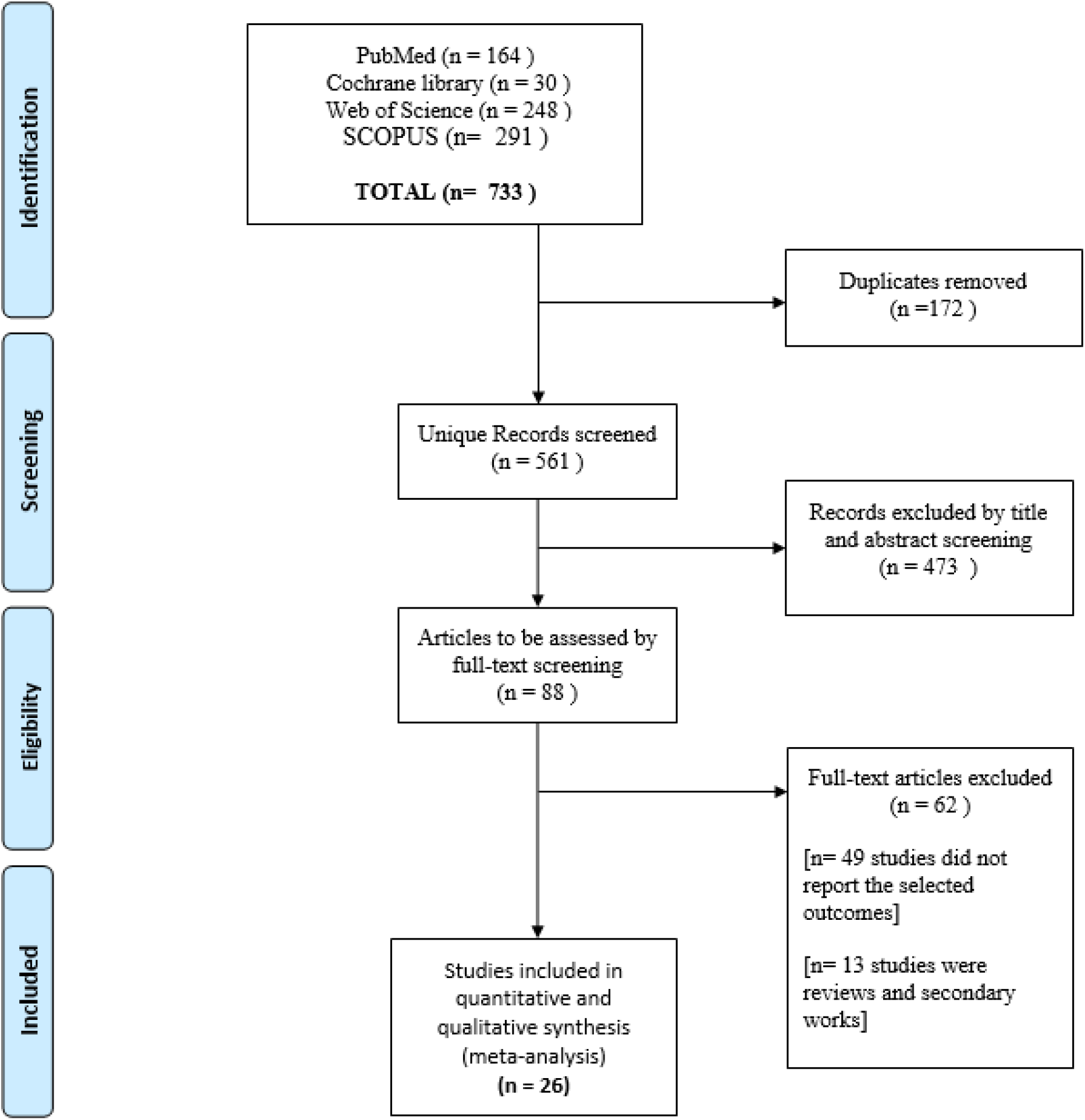
PRISMA diagram for our literature search

**Table 1:**
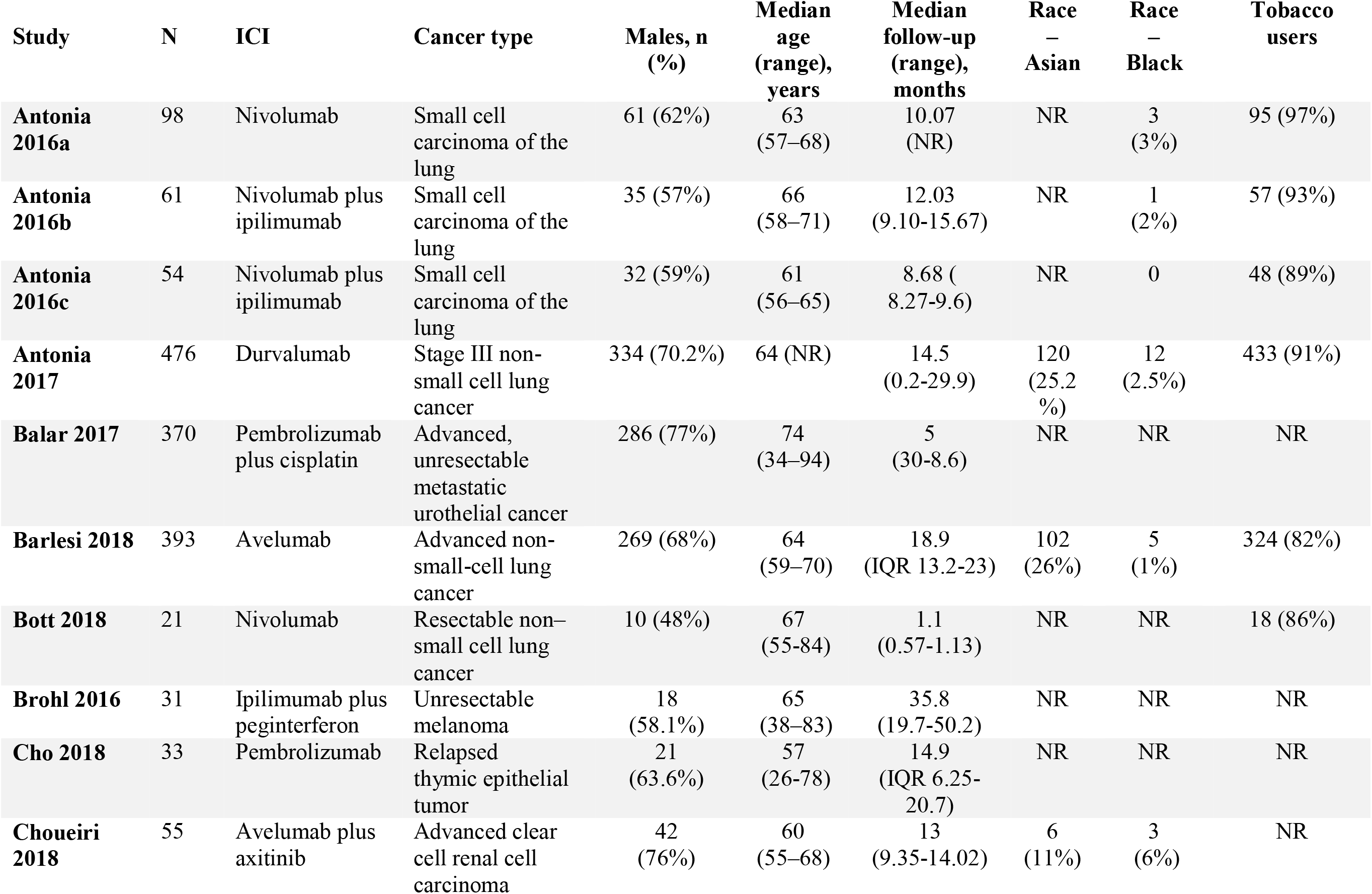
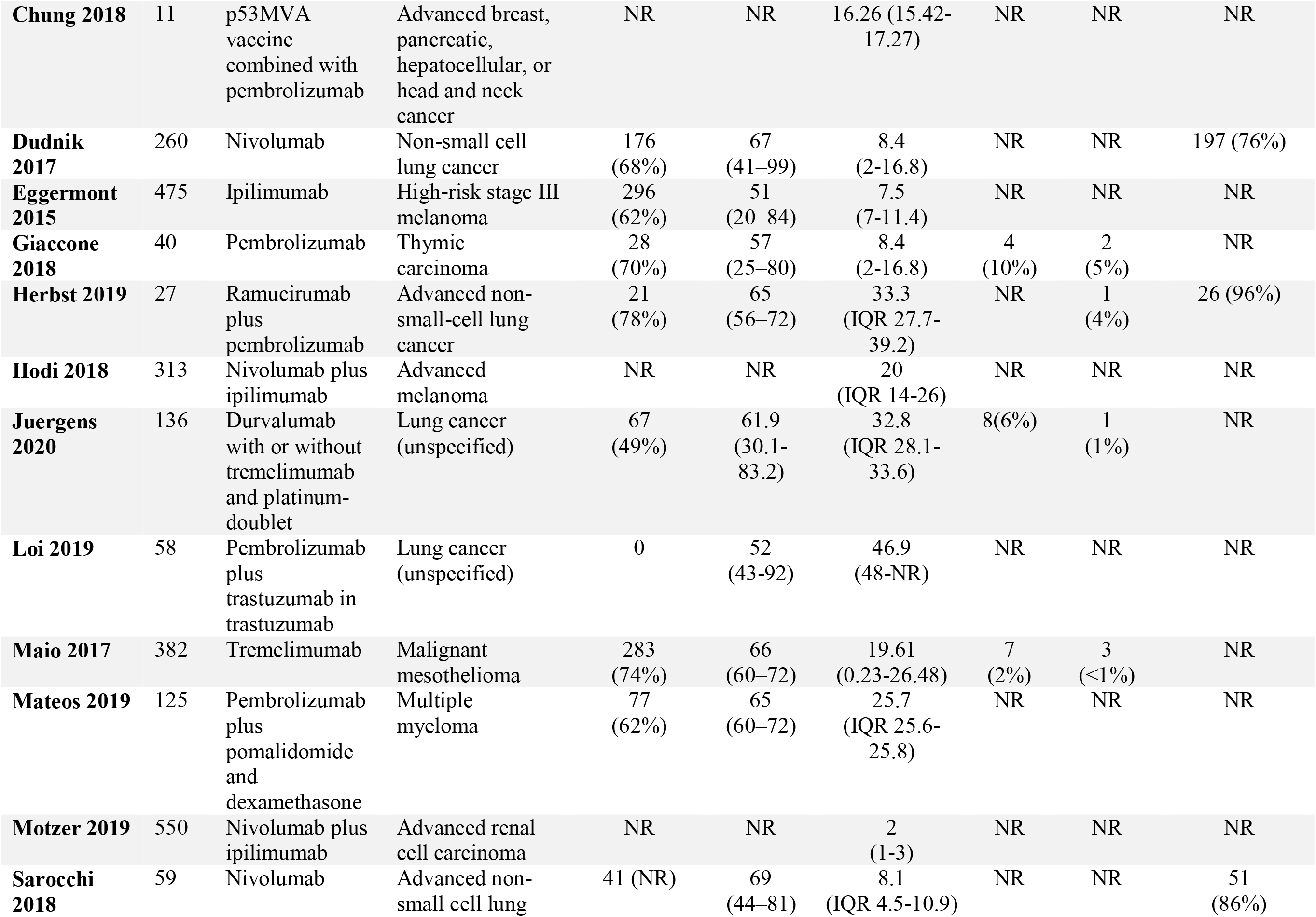
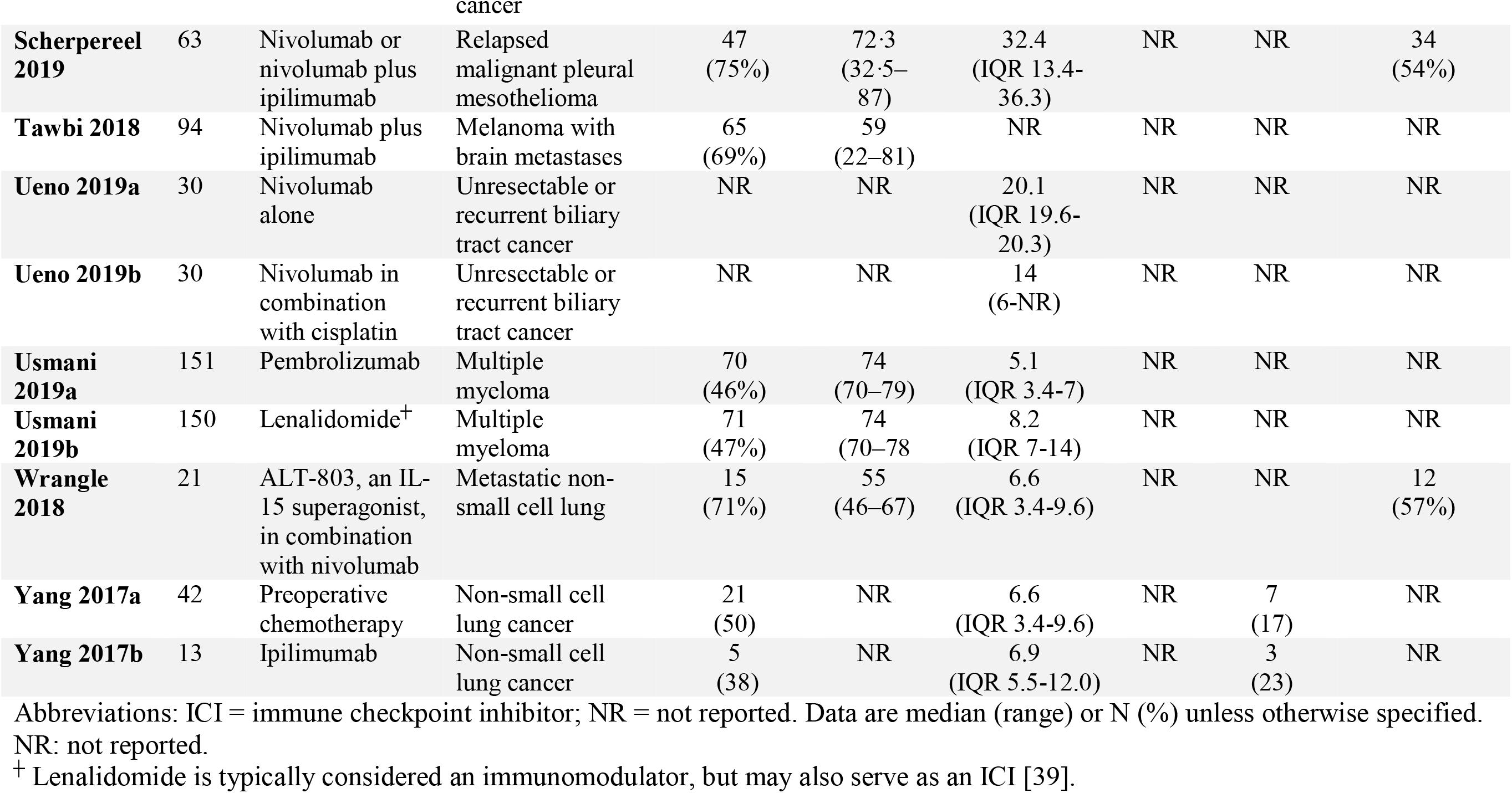
Summary of baseline characteristics of included studies.

| Study | N | ICI | Cancer type | Males, n (%) | Median age (range), years | Median follow-up (range), months | Race – Asian | Race – Black | Tobacco users |
| --- | --- | --- | --- | --- | --- | --- | --- | --- | --- |
| **Antonia 2016a** | 98 | Nivolumab | Small cell carcinoma of the lung | 61 (62%) | 63 (57–68) | 10.07 (NR) | NR | 3 (3%) | 95 (97%) |
| **Antonia 2016b** | 61 | Nivolumab plus ipilimumab | Small cell carcinoma of the lung | 35 (57%) | 66 (58–71) | 12.03 (9.10-15.67) | NR | 1 (2%) | 57 (93%) |
| **Antonia 2016c** | 54 | Nivolumab plus ipilimumab | Small cell carcinoma of the lung | 32 (59%) | 61 (56–65) | 8.68 (8.27-9.6) | NR | 0 | 48 (89%) |
| **Antonia 2017** | 476 | Durvalumab | Stage III non-small cell lung cancer | 334 (70.2%) | 64 (NR) | 14.5 (0.2-29.9) | 120 (25.2%) | 12 (2.5%) | 433 (91%) |
| **Balar 2017** | 370 | Pembrolizumab plus cisplatin | Advanced, unresectable metastatic urothelial cancer | 286 (77%) | 74 (34–94) | 5 (30-8.6) | NR | NR | NR |
| **Barlesi 2018** | 393 | Avelumab | Advanced non-small-cell lung cancer | 269 (68%) | 64 (59–70) | 18.9 (IQR 13.2-23) | 102 (26%) | 5 (1%) | 324 (82%) |
| **Bott 2018** | 21 | Nivolumab | Resectable non-small cell lung cancer | 10 (48%) | 67 (55-84) | 1.1 (0.57-1.13) | NR | NR | 18 (86%) |
| **Brohl 2016** | 31 | Ipilimumab plus peginterferon | Unresectable melanoma | 18 (58.1%) | 65 (38–83) | 35.8 (19.7-50.2) | NR | NR | NR |
| **Cho 2018** | 33 | Pembrolizumab | Relapsed thymic epithelial tumor | 21 (63.6%) | 57 (26-78) | 14.9 (IQR 6.25-20.7) | NR | NR | NR |
| **Choueiri 2018** | 55 | Avelumab plus axitinib | Advanced clear cell renal cell carcinoma | 42 (76%) | 60 (55–68) | 13 (9.35-14.02) | 6 (11%) | 3 (6%) | NR |
| **Chung 2018** | 11 | p53MVA vaccine combined with pembrolizumab | Advanced breast, pancreatic, hepatocellular, or head and neck cancer | NR | NR | 16.26 (15.42-17.27) | NR | NR | NR |
| **Dudnik 2017** | 260 | Nivolumab | Non-small cell lung cancer | 176 (68%) | 67 (41–99) | 8.4 (2-16.8) | NR | NR | 197 (76%) |
| **Eggermont 2015** | 475 | Ipilimumab | High-risk stage III melanoma | 296 (62%) | 51 (20–84) | 7.5 (7-11.4) | NR | NR | NR |
| **Giaccone 2018** | 40 | Pembrolizumab | Thymic carcinoma | 28 (70%) | 57 (25–80) | 8.4 (2-16.8) | 4 (10%) | 2 (5%) | NR |
| **Herbst 2019** | 27 | Ramucirumab plus pembrolizumab | Advanced non-small-cell lung cancer | 21 (78%) | 65 (56–72) | 33.3 (IQR 27.7-39.2) | NR | 1 (4%) | 26 (96%) |
| **Hodi 2018** | 313 | Nivolumab plus ipilimumab | Advanced melanoma | NR | NR | 20 (IQR 14-26) | NR | NR | NR |
| **Juergens 2020** | 136 | Durvalumab with or without tremelimumab and platinum-doublet | Lung cancer (unspecified) | 67 (49%) | 61.9 (30.1-83.2) | 32.8 (IQR 28.1-33.6) | 8(6%) | 1 (1%) | NR |
| **Loi 2019** | 58 | Pembrolizumab plus trastuzumab in trastuzumab | Lung cancer (unspecified) | 0 | 52 (43-92) | 46.9 (48-NR) | NR | NR | NR |
| **Maio 2017** | 382 | Tremelimumab | Malignant mesothelioma | 283 (74%) | 66 (60–72) | 19.61 (0.23-26.48) | 7 (2%) | 3 (<1%) | NR |
| **Mateos 2019** | 125 | Pembrolizumab plus pomalidomide and dexamethasone | Multiple myeloma | 77 (62%) | 65 (60–72) | 25.7 (IQR 25.6-25.8) | NR | NR | NR |
| **Motzer 2019** | 550 | Nivolumab plus ipilimumab | Advanced renal cell carcinoma | NR | NR | 2 (1-3) | NR | NR | NR |
| **Sarocchi 2018** | 59 | Nivolumab | Advanced non-small cell lung | 41 (NR) | 69 (44–81) | 8.1 (IQR 4.5-10.9) | NR | NR | 51 (86%) |
| **Scherpereel 2019** | 63 | Nivolumab or nivolumab plus ipilimumab | Relapsed malignant pleural mesothelioma | 47<br>(75%) | 72.3<br>(32.5–87) | 32.4<br>(IQR 13.4–36.3) | NR | NR | 34<br>(54%) |
| **Tawbi 2018** | 94 | Nivolumab plus ipilimumab | Melanoma with brain metastases | 65<br>(69%) | 59<br>(22–81) | NR | NR | NR | NR |
| **Ueno 2019a** | 30 | Nivolumab alone | Unresectable or recurrent biliary tract cancer | NR | NR | 20.1<br>(IQR 19.6–20.3) | NR | NR | NR |
| **Ueno 2019b** | 30 | Nivolumab in combination with cisplatin | Unresectable or recurrent biliary tract cancer | NR | NR | 14<br>(6–NR) | NR | NR | NR |
| **Usmani 2019a** | 151 | Pembrolizumab | Multiple myeloma | 70<br>(46%) | 74<br>(70–79) | 5.1<br>(IQR 3.4–7) | NR | NR | NR |
| **Usmani 2019b** | 150 | Lenalidomide<sup>†</sup> | Multiple myeloma | 71<br>(47%) | 74<br>(70–78) | 8.2<br>(IQR 7–14) | NR | NR | NR |
| **Wrangle 2018** | 21 | ALT-803, an IL-15 superagonist, in combination with nivolumab | Metastatic non-small cell lung | 15<br>(71%) | 55<br>(46–67) | 6.6<br>(IQR 3.4–9.6) | NR | NR | 12<br>(57%) |
| **Yang 2017a** | 42 | Preoperative chemotherapy | Non-small cell lung cancer | 21<br>(50) | NR | 6.6<br>(IQR 3.4–9.6) | NR | 7<br>(17) | NR |
| **Yang 2017b** | 13 | Ipilimumab | Non-small cell lung cancer | 5<br>(38) | NR | 6.9<br>(IQR 5.5–12.0) | NR | 3<br>(23) | NR |

Abbreviations: ICI = immune checkpoint inhibitor; NR = not reported. Data are median (range) or N (%) unless otherwise specified. NR: not reported.
<sup>†</sup> Lenalidomide is typically considered an immunomodulator, but may also serve as an ICI [39].

#### Data extraction

We used a data extraction form specifically designed for this study. Three main categories of data were extracted. The first category included baseline data about the study participants, such as patients’ age, gender, cancer type, and drug administered and dose. The second category included different outcome endpoints for analysis (any reported cardiovascular adverse event). The third category involved data used to assess the risk of bias among the included studies.

#### Quality and risk of bias assessment

This systematic review and meta-analysis were conducted in accordance with the principles of the Grading of Recommendations, Assessment, Development, and Evaluations (GRADE). We included clinical trials only to ensure high-quality evidence. For assessment of the risk of bias, we used the Cochrane’s Risk of Bias tool (21).

#### Data synthesis

The extracted data were restricted to dichotomous outcomes, as all the data for the analysis are adverse events expressed as events/total. Using the OpenMeta[Analyst] Software, the intended scores were pooled as risk ratios (RR), and the presence of heterogeneity was assessed using two main tests (22), the I-square test (*I^2^*) and the P-value of the Chi-square test. The analysis is said to be heterogeneous if values of *I^2^*>50% and p < 0.1 were present, according to the Cochrane Handbook (20). We performed the analysis of homogeneous data under a fixed-effects model, while heterogeneous data were analyzed under the random-effects model.

## Results

### Summary of included studies

We present the analysis of 4,622 cancer patients from 26 studies. Figure 1 presents a flow diagram of the number of studies at each stage of the study selction process. Males were slightly overrepresented as compared to females (2,420 [52.4%] versus 2,202 [47.6%]). The mean age was 63.7 years. Further details pertaining to study characteristics, cancer type, ICI administered, and demographic data are illustrated in Table 1.

### Results of risk of bias

The overall risk of bias was high among the included studies. Studies reported various data regarding randomization of patients, allocation concealment, blinding of participants and personnel, blinding of outcome assessors, attrition bias, and selective reporting. The risk of bias status is summarized inFigure 2.

**Figure 2.**
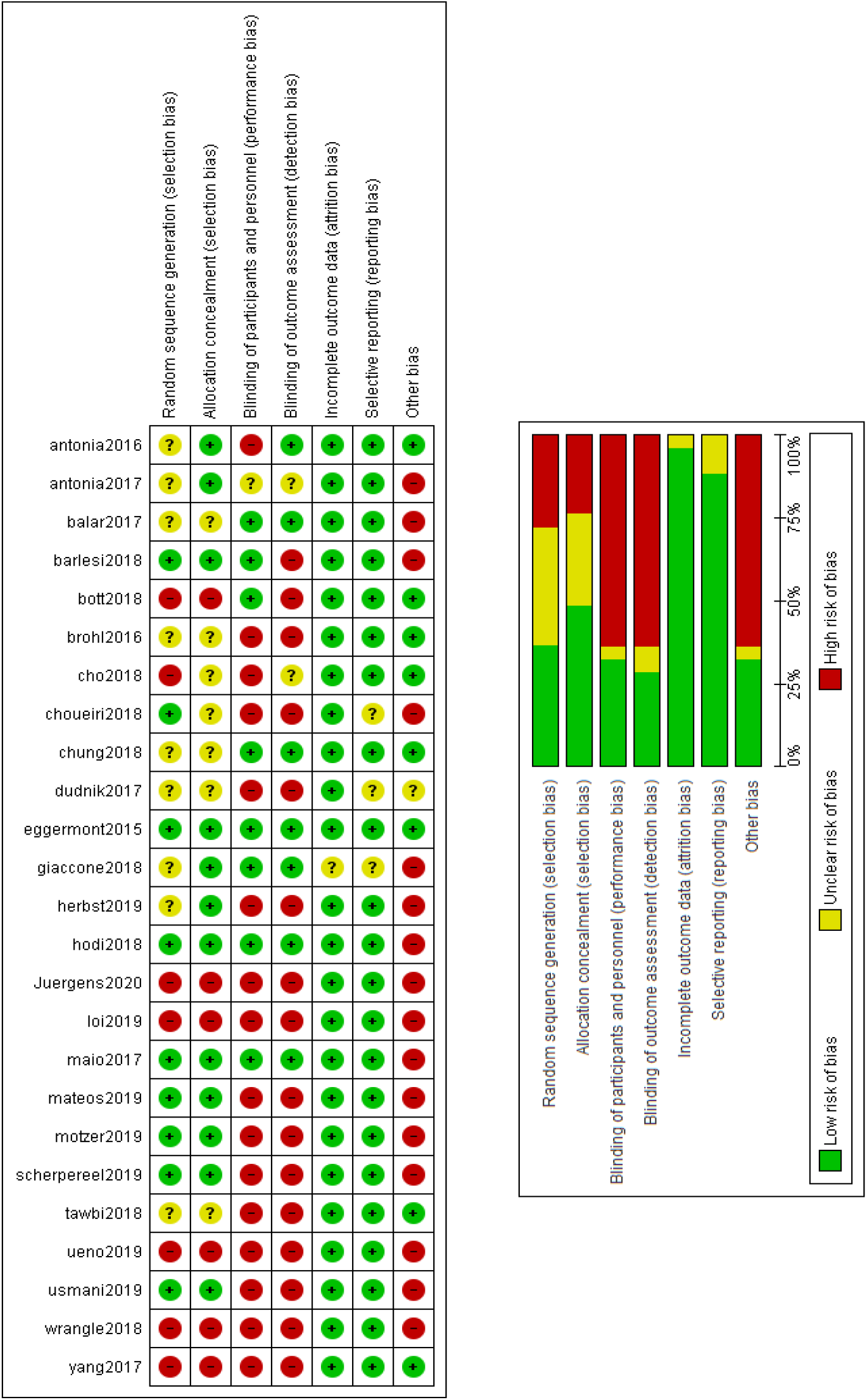
Results of risk of bias assessment among included trials.

### Results of analysis of outcomes

#### Incidence of myocarditis

Twelve studies reported the incidence of myocarditis as a cardiovascular irAE. The overall effect estimate showed that the incidence of myocarditis was 0.5%; the analysis was significant (95% CI [0.1%-0.9%]) and homogeneous (*I^2^* = 0%, P = 0.5).

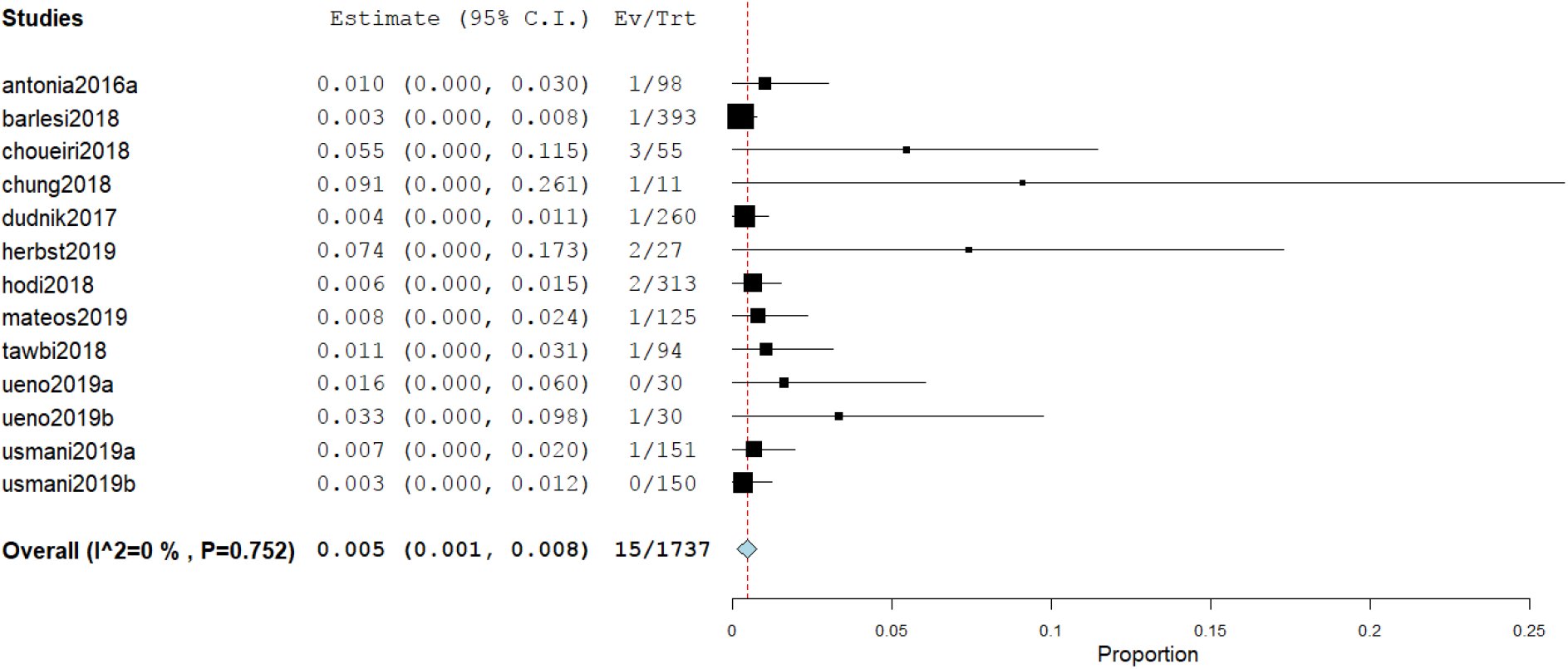

#### Incidence of pericardial effusion

Nine studies reported the incidence of pericardial effusion as a cardiovascular irAE. The overall effect estimate showed that the incidence of pericardial effusion was 0.5%; the analysis was significant (95% CI [0.1%-1.0%]) and homogeneous (*I^2^* = 36.7%, P = 0.1).

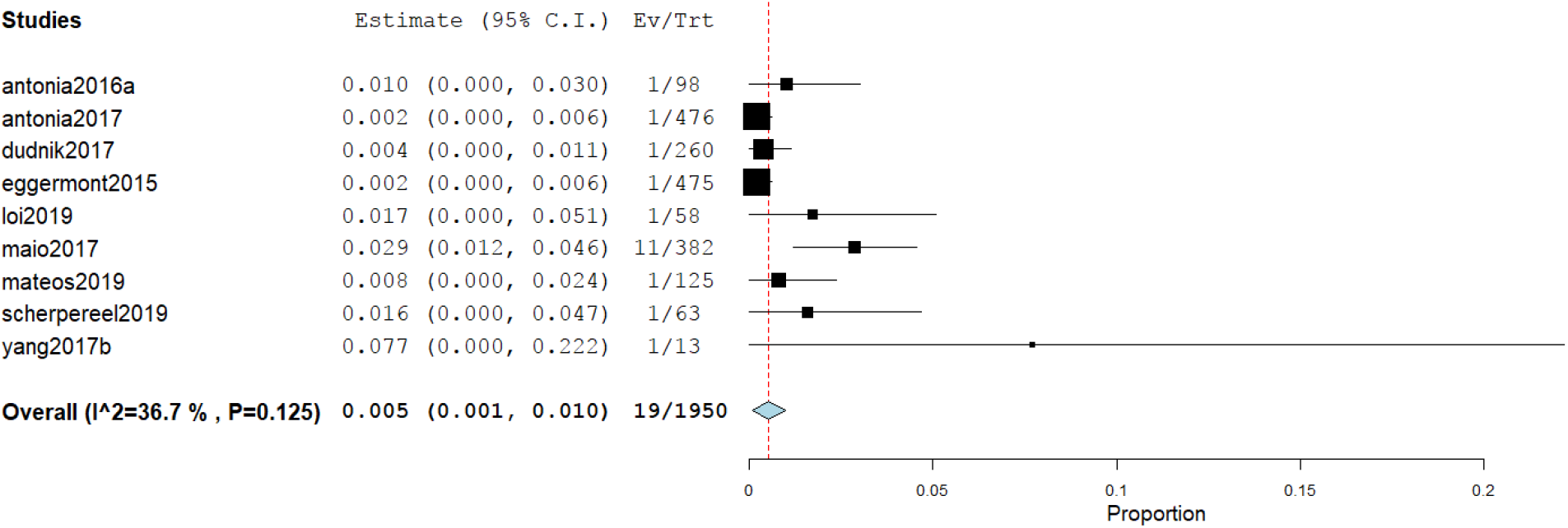

#### Incidence of heart failure

Seven studies reported the incidence of heart failure as a cardiovascular irAE. The overall effect estimate showed that the incidence of heart failure was 0.3%; the analysis was homogeneous (*I^2^* = 0%, P = 0.1) but not significant (95% CI [0.0%-0.5%]).

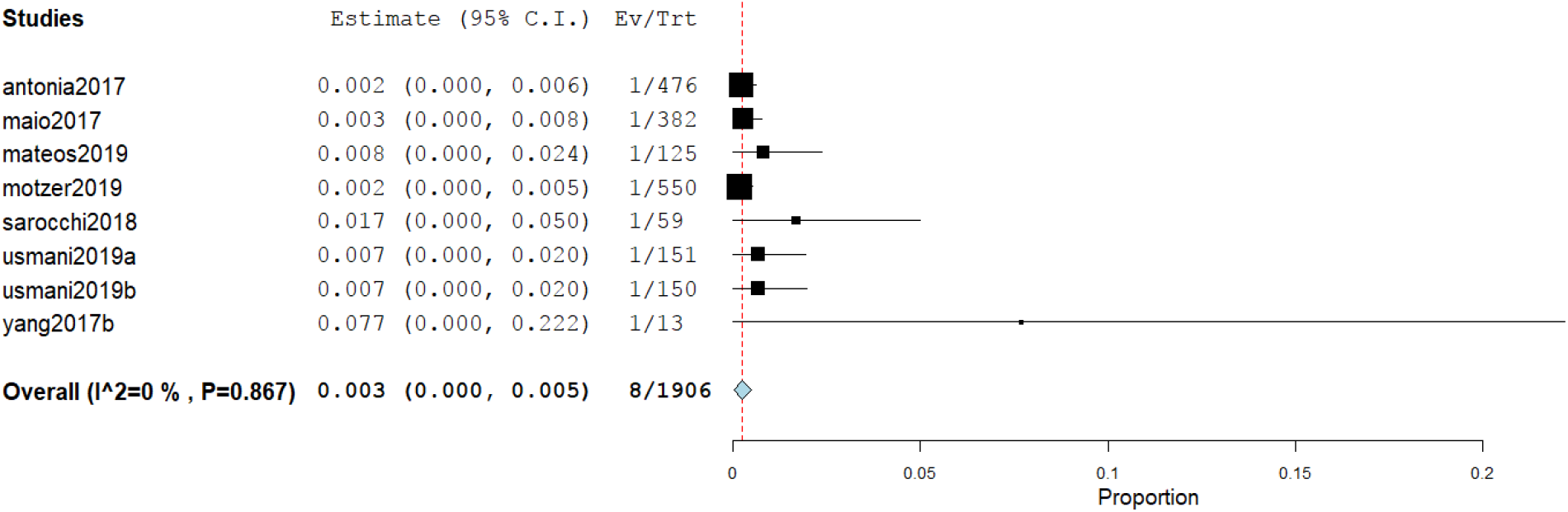

#### Incidence of cardiomyopathy

Five studies reported the incidence of cardiomyopathy as a cardiovascular irAE. The overall effect estimate showed that the incidence of cardiomyopathy was 0.3%; the analysis was homogeneous (*I^2^* = 0%, P = 0.6) but not significant (95% CI [-0.1%-0.6%]).

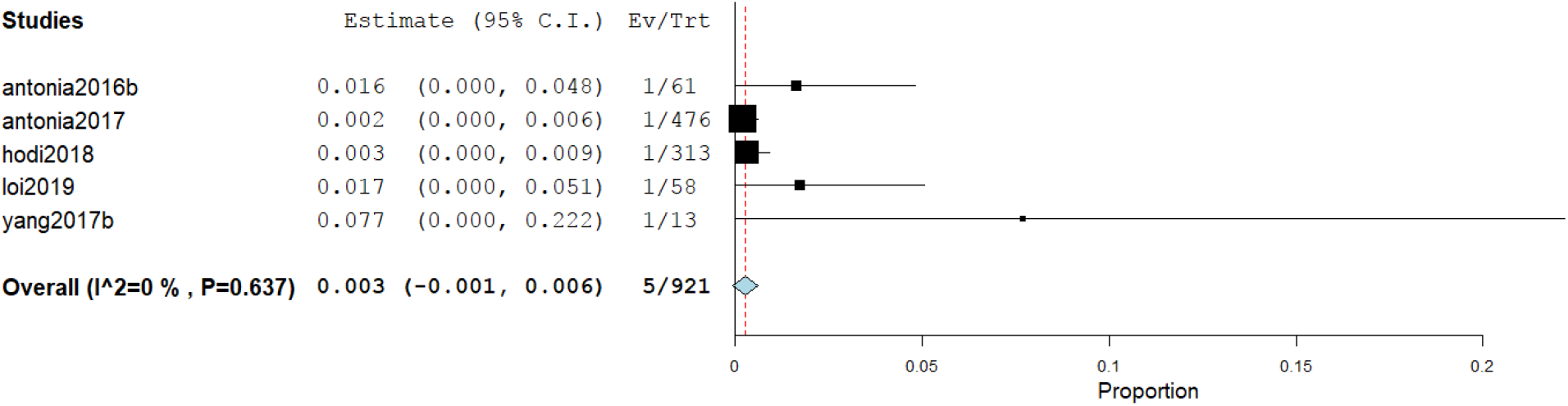

#### Incidence of atrial fibrillation

Four studies reported the incidence of atrial fibrillation as a cardiovascular irAE. The overall effect estimate showed that the incidence of atrial fibrillation was 7.6%.; the analysis was significant (95% CI [1.0%-14.1%]) and heterogeneous (*I^2^* = 0%, P = 0.6). Using Cochrane’s leave-one-out method, we solved the heterogeneity by excluding one study (Bott et al). Homogeneous results revealed an incidence rate of atrial fibrillation of 4.6%. The results were not significant (95% CI [-0.2%-9.4%]).

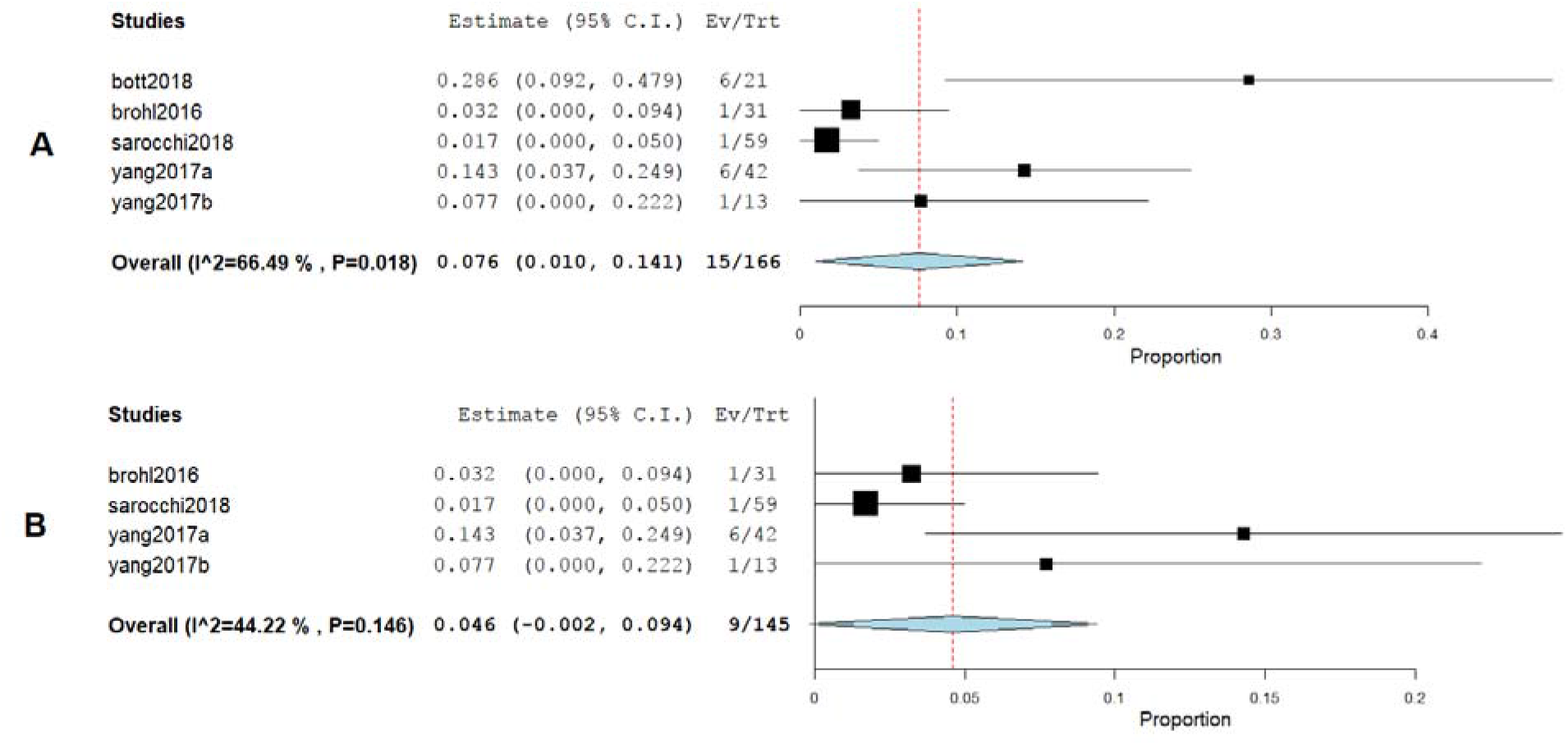

#### Incidence of myocardial infarction

Six studies reported the incidence of myocardial infarction as a cardiovascular irAE. The overall effect estimate showed that the incidence of MI was 0.4%; the analysis was homogeneous (*I^2^* = 0%, P = 0.1) but not significant (95% CI [0.0%-0.7%]).

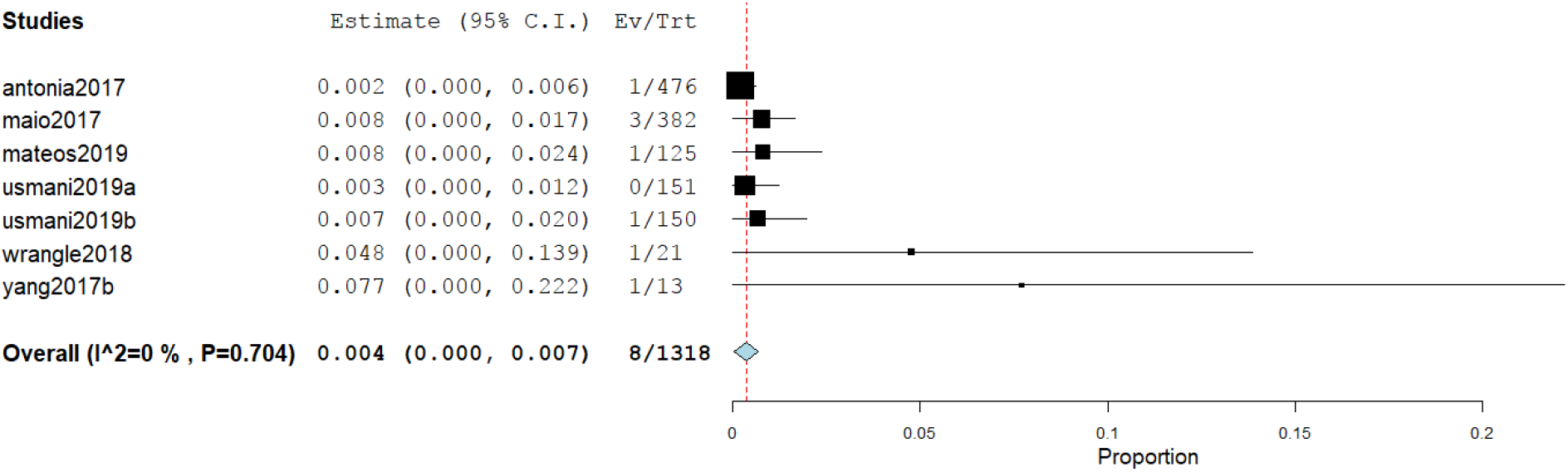

#### Incidence of cardiac arrest

Four studies reported the incidence of cardiac arrest as a cardiovascular irAE. The overall effect estimate showed that the incidence of cardiac arrest was 0.4%; the analysis was significant (95% CI [0.1%-0.8%]) and homogeneous (*I^2^* = 0%, P = 0.6)

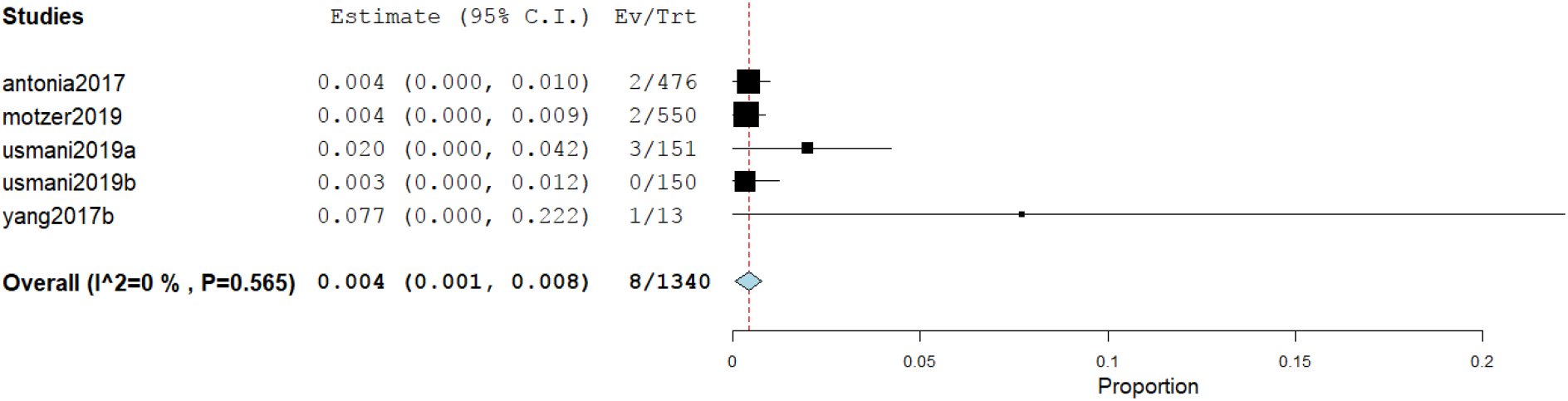

## Discussion

Cardiotoxicity is a rare but potentially fatal adverse effect of ICI therapy. The incidence of cardiovascular irAEs remains to be established (23). Our meta-analysis of 26 studies including a total of 4,622 ICI-treated cancer patients showed that 0.5% of cancer patients treated with ICIs developed myocarditis, 0.3% developed heart failure, and 7.6% developed atrial fibrillation. In addition, pericardial effusion occurred in 0.5% of patients, cardiomyopathy in 0.3% of patients, myocardial infarction in 0.4% of patients, and cardiac arrest in 0.4% of patients. These results are relatively consistent as evidenced by the low level of statistical heterogeneity.

The underlying pathogenesis of cardiovascular irAEs has yet to be fully elucidated. However, several mechanisms have been proposed. The most frequently postulated mechanism underlying myocarditis is that T-lymphocytes could target an antigen common to both neoplastic tissue and the heart. Indeed, in a recent report by Johnson et al., the authors described a common high-frequency T-lymphocyte sequence found in both tumor and cardiac muscles (18). Preclinical studies of mouse models have also shown that PD-1 and CTLA-4 deficiency is associated with myocarditis. The deletion of the PD-1 and CTLA-4 axes induces autoimmune myocarditis, indicating that the PD-1/PD-L1 interaction and CLTA-4 play important roles in protecting against T-lymphocyte-mediated inflammation (24–26). Injury usually occurs within first three months of initiating ICI; however, late presentation is not uncommon (27, 28).

T-lymphocyte-mediated inflammation may also be implicated in the pathogenesis of ICI-related atrial fibrillation. In one recent case report, histopathologic analysis of a patient with atrial fibrillation displayed patchy infiltrations of lymphocytes in the sinoatrial and atrioventricular nodes (18); this suggests that T-lymphocytes are intricately involved in the development of atrial fibrillation and other ICI-induced conduction disorders.

T-lymphocyte-related inflammatory processes are also suspected in pericardial effusion(29) and myocardial infarction (30). Lyon et al. suggested that the development of ICI-induced myocardial infarction could be due to the activation of an inflammatory reaction that triggers atherosclerotic coronary plaque formation and acute infarction. Conversely, Nykl et al. argued that the PD-1 inhibitory effect of ICIs leads to coronary vasospasm and ST-segment elevation. The mechanism by which coronary vasospasm develops is unclear but could be associated with systemic inflammatory response syndrome (21).

The incidence of cardiovascular irAEs is affected by many risk factors. Patients treated with combination therapy were more susceptible to cardiac complications as compared to those treated with ICI monotherapy (27). In addition, male patients are at higher risk of developing cardiovascular irAEs. A retrospective analysis showed that 77% of cases with ICI-related cardiac toxicity were males (26). In addition, another multicenter study found that 23 out of 35 irAEs (71%) occurred in male patients (32). However, data is limited and based on retrospective analyses of a small number of cases (65 cases). Furthermore, concomitant cardiovascular disease is a potential risk factor for cardiovascular irAEs (33).

Cardiovascular irAEs are classified into four grades by the Society for Immunotherapy of Cancer (34). The management of patients with cardiovascular irAEs differs based on the grade and severity of the symptoms. Grade I is usually asymptomatic and requires neither treatment nor discontinuation of immunotherapy. Grade II is characterized by mild cardiac symptoms that should be controlled by holding cancer immunotherapy and management of the coexisting cardiac disease and its risk factors. Grade III cardiovascular symptoms are significant and require the withdrawal of ICI therapy as well as urgent initiation of high-dose prednisone (1–2 mg/kg). Grade IV cardiovascular irAEs are life-threatening conditions characterized by decompensated cardiac function with moderate-to-severe symptoms; corticosteroid therapy is the first-line treatment. The addition of intravenous immunoglobulins, infliximab, or anti-thymocyte globulin should be considered as second-line treatments for patients with grade IV cardiovascular irAEs (10, 34).

Long-term data regarding the prognosis of patients with cardiovascular irAEs are limited. However, the available findings suggest a high fatality rate. In a systematic review that included 99 patients with cardiovascular irAEs, the fatality rate was 35% (28, 35). In addition, observational studies report a 50% rate of major adverse cardiac events in ICI-associated myocarditis, which is significantly higher than that of non-ICI-related myocarditis (36, 37).

This study represents an attempt to estimate the overall incidence of cardiovascular irAEs in cancer patients receiving ICI therapy. The quality of the included studies ranged from low to moderate according to the Cochrane Risk of Bias Assessment tool (21). The main limitation of our analysis is that the included studies were not primarily designed to investigate the incidence of ICI-induced cardiac adverse events. In addition, there was a high risk of bias resulting from the difficultly in blinding and randomization of some studies. The definitions to determine adverse events were slightly different across all studies. We did not consider medication dose, which may influence the severity of adverse effects. It is also important to note that malignancy in and of itself is a risk factor for coronary artery disease and other cardiovascular comorbidities and hence it is difficult to differentiate a concomitant cardiovascular irAE (38). It is therefore reasonable to perform cardiovascular magnetic resonance to distinguish a pre-existing cardiovascular disease from a cardiovascular irAE (36, 38). Nevertheless, we believe this analysis provides a valuable framework for further studies on ICI-associated cardiovascular events.

## Conclusion

Cardiovascular irAEs are rare but potentially life-threatening complications that can occur in patients receiving ICI therapy. Our analysis revealed that the most frequent ICI-associated adverse events are atrial fibrillation, myocarditis, and pericardial effusion. Risk factors for cardiovascular irAEs include treatment with combination immunotherapy, male sex, and a history of cardiac disease. Data on the prognosis of cardiac irAEs are limited. Ongoing post-market surveillance is therefore imperative to characterize long-term risks and improve outcomes among patients receiving ICIs.

## Data Availability

Data is available from the authors upon reasonable request.

## Author Contributions

DA conceived the study hypothesis. NN and DA designed the study and performed the systematic search, study selection and data extraction. NN analyzed the data. All authors contributed to the interpretation of the data, writing and critical editing of the manuscript.

## Protocol Registration

None

## Funding

The study was not supported by any funding sources

## Conflicts of interest

None declared.

## Notes

### Competing Interest Statement

The authors have declared no competing interest.

### Funding Statement

No external funding was received.

### Author Declarations

This systematic review and meta-analysis is exempt from Institutional Review Board oversight.

